# Remdesivir for the treatment of COVID-19 disease: A retrospective comparative study of patients treated with and without Remdesivir

**DOI:** 10.1101/2021.07.15.21260600

**Authors:** Surabhi Madan, Amit Patel, Kartikae Sharan, Shayon Ghosh, Vishnu Venugopal, Nitesh Shah, Bhagyesh Shah, Vipul Thakkar, Rashmi Chovatiya, Hardik Shah, Pradip Dabhi, Minesh Patel, Bhowmik Meghnathi, Vineet Sankhla, Vipul Kapoor, Tejas Patel, Maulik Soni, Nirav Bapat, Kaivan Shah, Ritanshu Chandarana, Parloop Bhatt, Manish Rana

**Author notes:** Corresponding author: Dr Surabhi Madan, Consultant, Department of Infectious Disease, Address: CIMS Hospital, Science City Road, Sola, Ahmedabad, Gujarat, India.

## Abstract

**Background:** Remdesivir (RDV) in coronavirus disease 2019 (COVID-19) has been found to be beneficial in patients with severe disease; however, its role in mild-moderate disease and its optimal timing need to be identified.

**Objective:** To assess the course of illness and final outcome in patients who received RDV at various stages of illness, and compare it to the non-RDV group.

**Methods:** This is a retrospective data analysis of 1262 COVID-19 patients hospitalized from May5, 2020 to August 31, 2020. The primary outcomes were progression to mechanical ventilation (MV) or death. Kaplan Meier survival analysis and log rank test were used for evaluating primary outcomes.

**Results:** 398 patients comprised the RDV group and 260 patients comprised the non-RDV group. 2/3^rd^ of patients were above 50 years of age in both the groups and 3/4^th^ patients were male. Mortality rate was 5.8% in RDV group (10.4% in non-RDV group). Mortality rate was 3.6%, 4% and 16.7% when RDV was started within 5 days, 5 to 10 days and after 10 days of symptom onset respectively. Fewer patients in RDV group progressed to MV (4.0% v/s 8.2%). Earlier discharge occurred in RDV group. Use of supplemental oxygen was observed in 44.7% patients in RDV group (54.2% in non-RDV group). No significant adverse events were observed with RDV. Survival analysis showed that probability of event (death) was significant for patients with hypertension (HT) and/or diabetes mellitus (DM) in RDV group.

**Conclusion:** Early initiation of RDV is associated with shorter hospital stay, lower mortality as well as reduced need for supplemental oxygen and mechanical ventilation.

## Introduction

Since its identification in December 2019, COVID-19 still continues to be a major global health and socioeconomic burden (1). Several therapeutic agents targeting the different aspects and phases of the disease, including various antiviral agents, have been evaluated for the treatment of COVID-19 (2).

RDV, a prodrug of an adenosine analogue that inhibits viral RNA-dependent RNA polymerase, was identified early as a promising drug for COVID-19 because of its ability to inhibit severe acute respiratory syndrome coronavirus 2 (SARS-CoV-2) in vitro (3). In October 2020, RDV received the U.S. FDA approval, however, the optimal role of RDV still remains uncertain and debatable (4). World Health Organization (WHO) does not suggest using it in hospitalized patients (5). Other guideline panels, including the Infectious Diseases Society of America and the National Institutes of Health, suggest using RDV in hospitalized patients who require supplemental oxygen (6, 7).

The objective of our study was to assess the course of illness and final outcome in patients who received RDV at various stages of illness, and compare it to those who were not administered RDV.

## Methods

### Study design and setting

This is a retrospective review and data analysis of 1262 COVID-19 patients hospitalized from May 5, 2020 to August 31, 2020 in a tertiary care private hospital of western India. The study was approved by the CIMS (Care Institute of Medical Sciences) hospital ethics committee (CTRI/2020/05/025247). The need for consent was waived off due to the nature of the study.

### Treatment protocol

We started admitting COVID-19 patients in the first week of May 2020. Though there were minor inter-consultant variations, a uniform treatment protocol was followed, which was based on the categorization of patients into mild, moderate, and severe categories, as defined by the Government of India; wherein mild disease is defined as the presence of upper respiratory infection without evidence of hypoxia, moderate disease is the presence of clinical features suggestive of pneumonia with SpO2<94% (range 90%-94%) on room air; and severe disease is clinical signs of pneumonia with SpO2<90% on room air (8).

The treatment protocol was updated regularly, as per the availability of new information regarding various drugs, which mainly included hydroxychloroquine (HCQ), azithromycin, RDV, anticoagulants and statins. All the patients received standard of care treatment, either HCQ (alone or in combination with azithromycin) in the early part of pandemic or RDV (once it got available, after which the use of HCQ became almost nil).

RDV was administered to all the patients who presented in the first two weeks of illness with fever for three or more days, or clinical signs suggestive of respiratory distress. Patients with high risk factors, like age 60 years or above, one or more comorbidity, along with radiological evidence of COVID-19 pneumonia, were treated with RDV even in the absence of fever, which varied as per the consultant’ s decision. RDV was not administered to asymptomatic patients. RDV was given at a dose of 200 mg loading dose on day 1, followed by a 100 mg maintenance dose, daily for a period of 5 days, though the duration could be prolonged up to 10 days at the treating doctor’ s discretion.

### Data collection

Demographic details, signs and symptoms, results of laboratory parameters, radiological investigations, ward and Intensive Care Unit (ICU) progress notes, and treatment and outcome details were obtained from the medical records files and hospital’ s intranet. Status of respiratory support on all the days during hospitalization was recorded. The clinical status of the patients was recorded on an eight-category ordinal scale, from the day of admission till discharge, as follows: 1. Discharged without oxygen support, 2. Discharged with oxygen support, 3. Hospitalized without fever and without oxygen support, 4. Hospitalized with fever and without oxygen support, 5 a. Hospitalized with oxygen support with Nasal Cannula (O2 via NC), 5 b. Hospitalized with oxygen support with mask (O2 via mask), 5 c. Hospitalized with oxygen support with non-rebreathing mask (O2 via NRBM), 6. Hospitalized with High Flow Nasal Cannula (HFNC) / Bilevel Positive Airway Pressure (BiPAP), 7. Hospitalized with Mechanical Ventilation (MV), 8. Death. The details were recorded in hard copy as well as in an electronic database.

### Outcomes

The primary outcome was progression to invasive mechanical ventilation or death. Secondary outcomes were discharge from the hospital (ordinal scale 1 and 2) by day 5, 7, 10 and 14; new oxygen use in patients who were not on supplemental oxygen on admission, progression to higher ordinal scales, and any adverse event including liver and kidney injury.

### Statistical analysis

Descriptive statistics were used for describing demographic and patient characteristics and were summarized as rates or percent for categorical variables, while median and 95% CI for median was used for the continuous variables. The change in continuous variables across the time point was assessed using the paired non-parametric Wilcoxon Signed-Rank Test. Kaplan Meier survival analysis was used for RDV and non-RDV groups and those with HT and/or DM, and log rank test was used for testing significance. Data were analyzed using IBM SPSS version 19.

## Results

1262 patients with a diagnosis of COVID-19 were admitted during the study period. RDV was available in our hospital since June 25, 2020. 398 of 1262 patients received RDV and comprised the intervention/ RDV group. The comparison group comprised of 527 patients admitted from May 5 till June 24 2020, during which period RDV was not available. Of these, 267 patients were excluded from the final analysis as they did not fulfill the criteria for RDV and were hospitalized for infection control and isolation purpose only, as per the national guidelines during early pandemic. 260 of 527 patients were included in the final analysis in the comparison / non-RDV group. 735 patients were admitted after June 25, 2020, of which 337 patients did not receive RDV, however the latter were not included in the comparison group as they did not fulfill the criteria for administration of RDV (Figure 1). There were 135 hospitalized patients in the ordinal scale 3 on the day of administration of RDV. Though no fever was recorded after hospitalization in these patients, they fulfilled the criteria for RDV as mentioned above.

**Figure 1:**
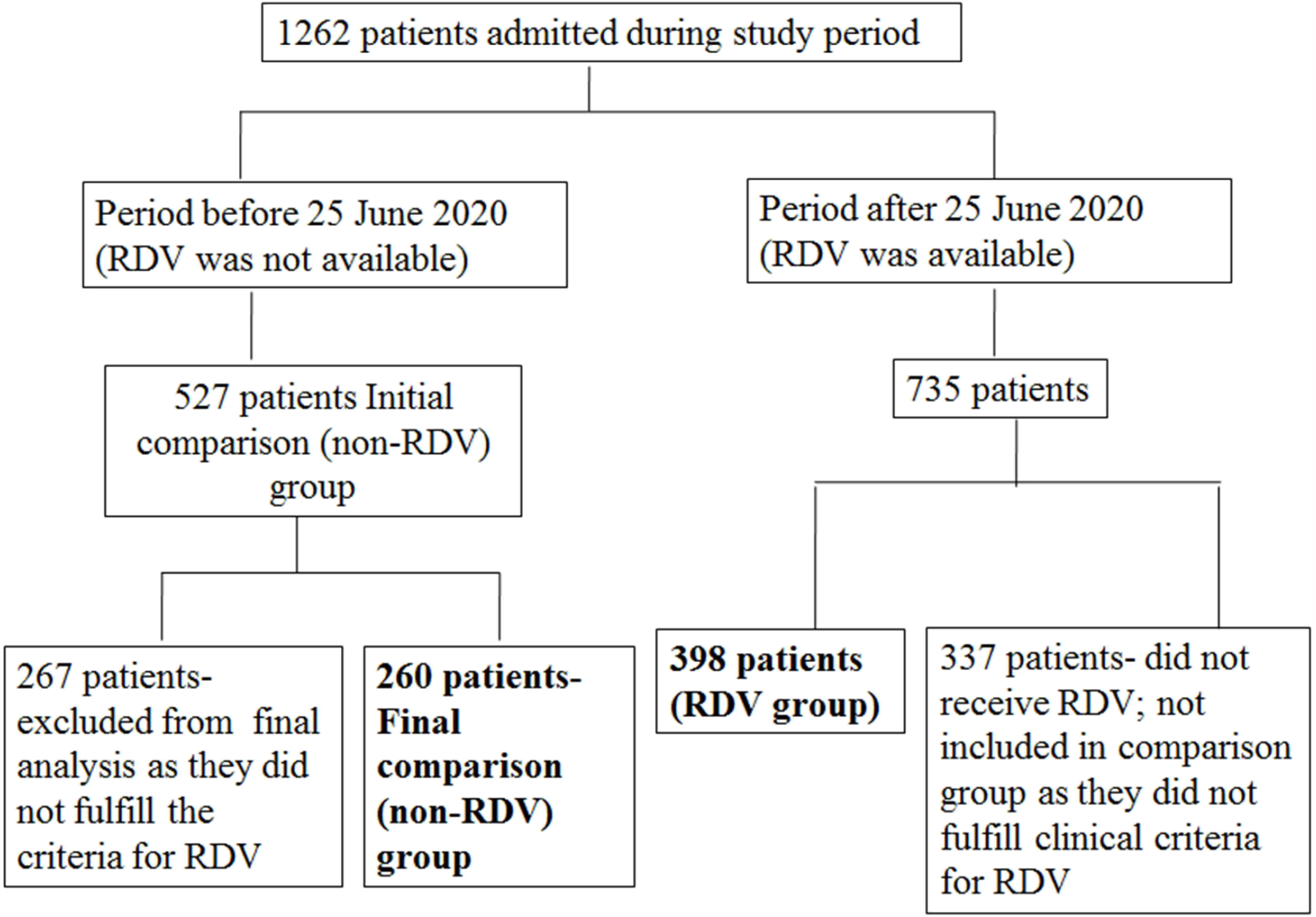
Flow chart depicting selection of patients

Majority of the patients were above 50 years of age (69.4% in RDV and 63.7% in non-RDV group). There were 78.1 % males in the RDV group and 73.8% males in the non-RDV group (table 1). HT and DM were the most common comorbidities in both the groups. RDV group had more number of patients with one or multiple comorbidities (table 2).

**Table 1:**
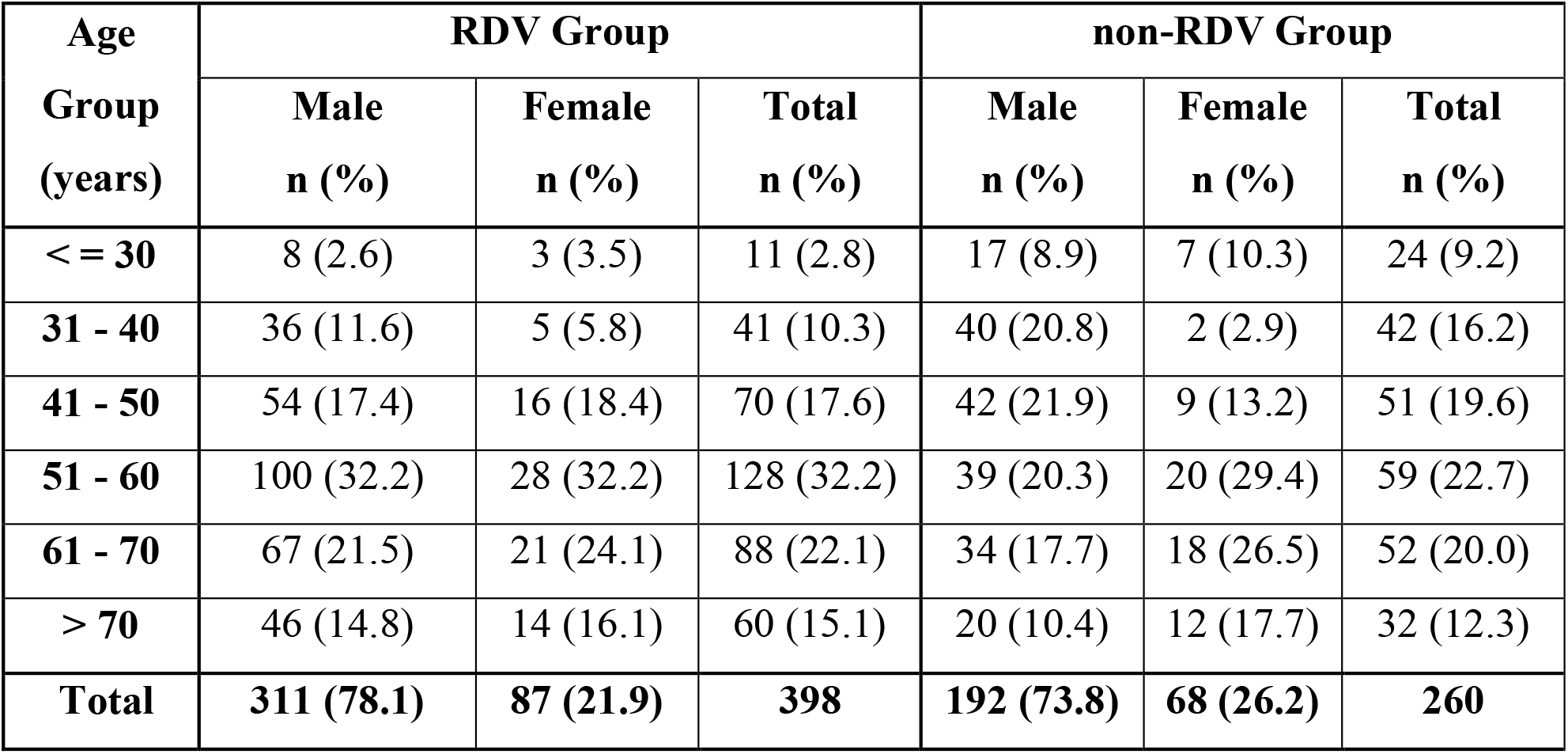
Age and sex distribution of patients in RDV and non-RDV group

**Table 2:** Comorbidities in RDV and non-RDV groups

| Comorbidity | RDV group (n=398) | non-RDV group (n=260) |
| --- | --- | --- |
|  | n (%) | n (%) |
| HT | 211 (53.0) | 129 (49.6) |
| DM | 171 (43.0) | 83 (31.9) |
| HT & DM | 122 (30.7) | 62 (23.8) |
| Obesity | 23 (5.8) | 12 (4.6) |
| Chronic cardiac disease | 68 (17.1) | 26 (10.0) |
| Chronic pulmonary disease | 18 (4.5) | 15 (5.8) |
| Chronic liver disease | 5 (1.3) | 2 (0.8) |
| Chronic kidney disease | 22 (5.5) | 6 (2.3) |
| Patient with no comorbidities | 89 (22.4) | 91 (35.0) |
| Patients with 1 or more comorbidities | 309 (77.6) | 169 (65.0) |

Table 3 depicts the relation between the timing of administration of RDV after the symptom onset and the disease outcome. Mortality rate was 3.6% when RDV was started within 5 days of symptom onset, as compared to16.7 %, when started after 10 days of symptom onset.

**Table 3:** Relation of timing of RDV to disease outcome

| Interval between symptom<br>onset and starting RDV | Outcome |  |  | Mortality |
| --- | --- | --- | --- | --- |
|  | Death | Alive | Total | Rate (%) |
| < 5 Days | 4 | 108 | 112 | 3.6 |
| 5 to 10 days | 9 | 217 | 226 | 4.0 |
| > 10 Days | 10 | 50 | 60 | 16.7 |
| Grand Total | 23 | 375 | 398 | 5.8 |

About 90% of the patients received RDV for a duration of 5 days. Death, before the completion of the standard 5 days treatment, and intermittent unavailability of the drug, were the main reasons for less than 5 days duration. About 5% patients received RDV for more than 5 days, which was done at the consultants’ discretion for critically ill patients. Mortality rate in patients who received RDV for 5 days was 3.7%, which was less than the mortality observed when RDV was given for less than and more than 5 days (35% and 13.6% respectively). Irrespective of the duration of RDV, mortality rate was highest when RDV was started after 10 days of symptom onset.

Tables 4 & 5 show the progressive changes in the ordinal scales during hospitalization in both the groups. Overall mortality rate during hospitalization was 5.8% in the RDV group as compared to 10.4% in the non-RDV group. In the RDV group, the proportion of patients discharged on day 5, 7, 10 and 14 of hospitalization were 40.7%, 67.1%, 83.9% and 89.2% respectively. In the non-RDV group, the respective rates were 26.9%, 49.6%, 70.4% and 81.2%.

**Table 4:** Change in ordinal scale of patients in RDV group

| RDV (n = 398 patients) |  |  |  |  |  |  |  |  |
| --- | --- | --- | --- | --- | --- | --- | --- | --- |
| Ordinal Scale | Category | Day of RDV | Day 3 | Day 5 | Day 7 | Day 10 | Day 14 | > Day 14 |
|  |  | N (%) | N (%) | N (%) | N (%) | N (%) | N (%) | N (%) |
| **1** | Discharged without oxygen support | 0 (0) | 6 (1.5) | 162 (40.7) | 267 (67.1) | 334 (83.9) | 354 (88.9) | 372 (93.4) |
| **2** | Discharged with oxygen support | 0 (0) | 0 (0) | 0 (0) | 0 (0) | 1 (0.3) | 1 (0.3) | 3 (0.8) |
| **3** | Hospitalized without fever and without oxygen support | 135 (33.9) | 243 (61.1) | 126 (31.7) | 48 (12.1) | 7 (1.8) | 1 (0.3) | 0 (0) |
| **4** | Hospitalized with fever and without oxygen support | 124 (31.2) | 18 (4.5) | 6 (1.5) | 4 (1.0) | 0 (0) | 0 (0) | 0 (0) |
| **5a** | Hospitalized with O2 via NC | 62 (15.6) | 62 (15.6) | 48 (12.1) | 32 (8) | 15 (3.8) | 7 (1.8) | 0 (0) |
| **5b** | Hospitalized with O2 via Mask | 20 (5.0) | 8 (2.0) | 7 (1.8) | 4 (1.0) | 3 (0.8) | 1 (0.3) | 0 (0) |
| **5c** | Hospitalized with NRBM | 24 (6.0) | 15 (3.8) | 9 (2.3) | 6 (1.5) | 4 (1.0) | 1 (0.3) | 0 (0) |
| **6** | HFNC / BiPaP | 26 (6.5) | 34 (8.5) | 28 (7.0) | 20 (5.0) | 12 (3.0) | 6 (1.5) | 0 (0) |
| **7** | MV | 7 (1.8) | 9 (2.3) | 7 (1.8) | 9 (2.3) | 8 (2.0) | 8 (2.0) | 0 (0) |
| **8** | Death | 0 (0) | 4 (1.0) | 5 (1.3) | 8 (2.0) | 14 (3.5) | 19 (4.8) | 23 (5.8) |

**Table 5:** Change in ordinal scale of patients in non-RDV group

| No Remdesivir (260 PTS) |  |  |  |  |  |  |  |  |
| --- | --- | --- | --- | --- | --- | --- | --- | --- |
| Ordinal Scale | Category | Day of Admission | Day 3 | Day 5 | Day 7 | Day 10 | Day 14 | > Day 14 |
|  |  | n (%) | n (%) | n (%) | n (%) | n (%) | n (%) | n (%) |
| **1** | Discharged without oxygen support | 0 (0) | 25 (9.6) | 70 (26.9) | 129 (49.6) | 183 (70.4) | 211 (81.2) | 233 (89.6) |
| **2** | Discharged with oxygen support | 0 (0) | 0 (0) | 0 (0) | 0 (0) | 0 (0) | 0 (0) | 0 (0) |
| **3** | Hospitalized without fever and without oxygen support | 20 (7.7) | 87 (33.5) | 82 (31.5) | 52 (20.0) | 20 (7.7) | 5 (1.9) | 0 (0) |
| **4** | Hospitalized with fever and without oxygen support | 154 (59.2) | 59 (22.7) | 24 (9.2) | 7 (2.7) | 2 (0.8) | 1 (0.4) | 0 (0) |
| **5a** | Hospitalized with O2 via NC | 41 (15.8) | 40 (15.4) | 35 (13.5) | 29 (11.2) | 17 (6.5) | 7 (2.7) | 0 (0) |
| **5b** | Hospitalized with O2 via Mask | 11 (4.2) | 6 (2.3) | 5 (1.9) | 4 (1.5) | 0 (0) | 0 (0) | 0 (0) |
| **5c** | Hospitalized with NRBM | 15 (5.8) | 5 (1.9) | 3 (1.2) | 3 (1.2) | 3 (1.2) | 1 (0.4) | 0 (0) |
| **6** | HFNC / BiPaP | 14 (5.4) | 27 (10.4) | 25 (9.6) | 15 (5.8) | 12 (4.6) | 8 (3.1) | 0 (0) |
| **7** | MV | 4 (1.5) | 8 (3.1) | 11 (4.2) | 11 (4.2) | 9 (3.5) | 9 (3.5) | 0 (0) |
| **8** | Death | 1 (0.4) | 3 (1.2)) | 5 (1.9) | 10 (3.8) | 14 (5.4) | 18 (6.9) | 27 (10.4) |

Defervescence occurred earlier in the RDV group. Patients with persistent fever on day 3 and 5 after the initiation of RDV were 4.5% and 1.5% respectively. These proportions in the non-RDV group were 22.7% and 9.2% respectively.

28.9% (115/398) patients in the RDV group needed low flow oxygen support (ordinal scale 5) as compared to 32.3% (84/260) in the non-RDV group. 10.1% (40/398) and 5.8% (23/398) patients required HFNC/BiPAP and MV respectively in the RDV group. The respective proportions in the non-RDV group were 11.5% (30/260) and 10.4% (27/260).

Steroids were administered to 73.4% of the patients in the RDV group as compared to 65.4% in the non-RDV group. However, in the subgroup where RDV was started within 5 days of symptom onset, 55.4% of the patients received steroids. 79.6% and 83.3% patients received steroids when RDV was started 5 to 10 days and more than 10 days after the symptom onset respectively.

Tocilizumab (TCZ) was administered to 22.9% of the patients in the RDV group as compared to 18.5% in the non-RDV group. In the subgroup where RDV was started within 5 days of symptom onset, 16.1% of the patients received TCZ. 20.8% and 43.3% patients received TCZ when RDV was started 5 to 10 days and more than 10 days after the symptom onset respectively.

Table 6 shows the number of patients who progressed to higher ordinal scales in both the groups. 55.3% patients in the RDV group did not require oxygen support as compared to 45.8% patients in the non-RDV group. 13.7% progressed from ordinal scale 4 to higher respiratory categories in RDV group while the same phenomenon was observed in 22% patients in the non-RDV group. Progression from ordinal scale 5 to scale 6 occurred in 15% of patients and to scale 7 occurred in 8.5% patients in RDV group. These rates were 17.9% and 13.4% respectively in the non-RDV group.

**Table 6:** Progression to higher ordinal scales in RDV and non-RDV group

| **Ordinal Scale** | **Respiratory category on day of starting RDV and day of admission in non-RDV group** | **Highest respiratory category during hospitalization** | **RDV Group n (%)** | **non-RDV group n (%)** |
| --- | --- | --- | --- | --- |
| 3 | Hospitalized without fever and without oxygen support | No fever and no O2 | 113 (83.7) | 0 (0) |
|  |  | O2 via NC | 14 (10.5) | 11 (55.0) |
|  |  | O2 via mask | 3 (2.2) | 2 (10.0) |
|  |  | O2 via NRBm | 1 (0.7) | 1 (5.0) |
|  |  | HFNC / BiPAP | 3 (2.2) | 4 (20.0) |
|  |  | MV | 1 (0.7) | 2 (10.0) |
|  |  | **Total** | **135** | **20** |
| 4 | Hospitalized with fever and without oxygen support | No fever and no O2 | 107 (86.3) | 119 (77.3) |
|  |  | O2 via NC | 15 (12.1) | 17 (11.0) |
|  |  | O2 via mask | 0 (0) | 6 (3.9) |
|  |  | O2 via NRBm | 1 (0.8) | 1 (0.7) |
|  |  | HFNC / BiPAP | 1 (0.8) | 5 (3.2) |
|  |  | MV | 0 (0) | 6 (3.9) |
|  |  | **Total** | **124** | **154** |
| 5 a | O2 via NC | O2 via NC | 43 (69.3) | 33 (80.5) |
|  |  | O2 via mask | 6 (9.7) | 3 (7.3) |
|  |  | O2 via NRBm | 3 (4.8) | 1 (2.4) |
|  |  | HFNC / BiPAP | 5 (8.1) | 4 (9.8) |
|  |  | MV | 5 (8.1) | 0 (0) |
|  |  | **Total** | **62** | **41** |
| 5 b | O2 via mask | O2 via mask | 12 (60.0) | 6 (54.5) |
|  |  | O2 via NRBm | 4 (20.0) | 0 (0) |
|  |  | HFNC / BiPAP | 1 (5.0) | 1 (9.1) |
|  |  | MV | 3 (15.0) | 4 (36.4) |
|  |  | **Total** | **20** | **11** |
| 5 c | O2 via NRBM | O2 via NRBM | 13 (54.1) | 3 (20.0) |
|  |  | HFNC / BiPAP | 10 (41.7) | 7 (46.7) |
|  |  | MV | 1 (4.2) | 5 (33.3) |
|  |  | **Total** | **24** | **15** |
| 6 | HFNC/BiPAP | HFNC / BiPAP | 20 (76.9) | 9 (64.3) |
|  |  | MV | 6 (23.1) | 5 (35.7) |
|  |  | **Total** | **26** | **14** |
| 7 | MV | MV | 7 (100.0) | 5 (100.0) |
|  |  | **Total** | **7** | **5** |
|  | **Grand Total** |  | **398** | **260** |

Among ordinal scale 6 in RDV group, 23% progressed to scale 7 while in the non-RDV group, 35% progressed to scale 7. When RDV was initiated while patient was already on MV, mortality was 100%. Progression to MV in the RDV group among ordinal scale 3 to 6 was 4.0% (16/398) while in the non-RDV group, it was 8.2% (22/260).

Mortality rate was lower in the RDV group compared to the non-RDV group in all the categories, except HFNC/BiPAP group (table 7).

**Table 7:**
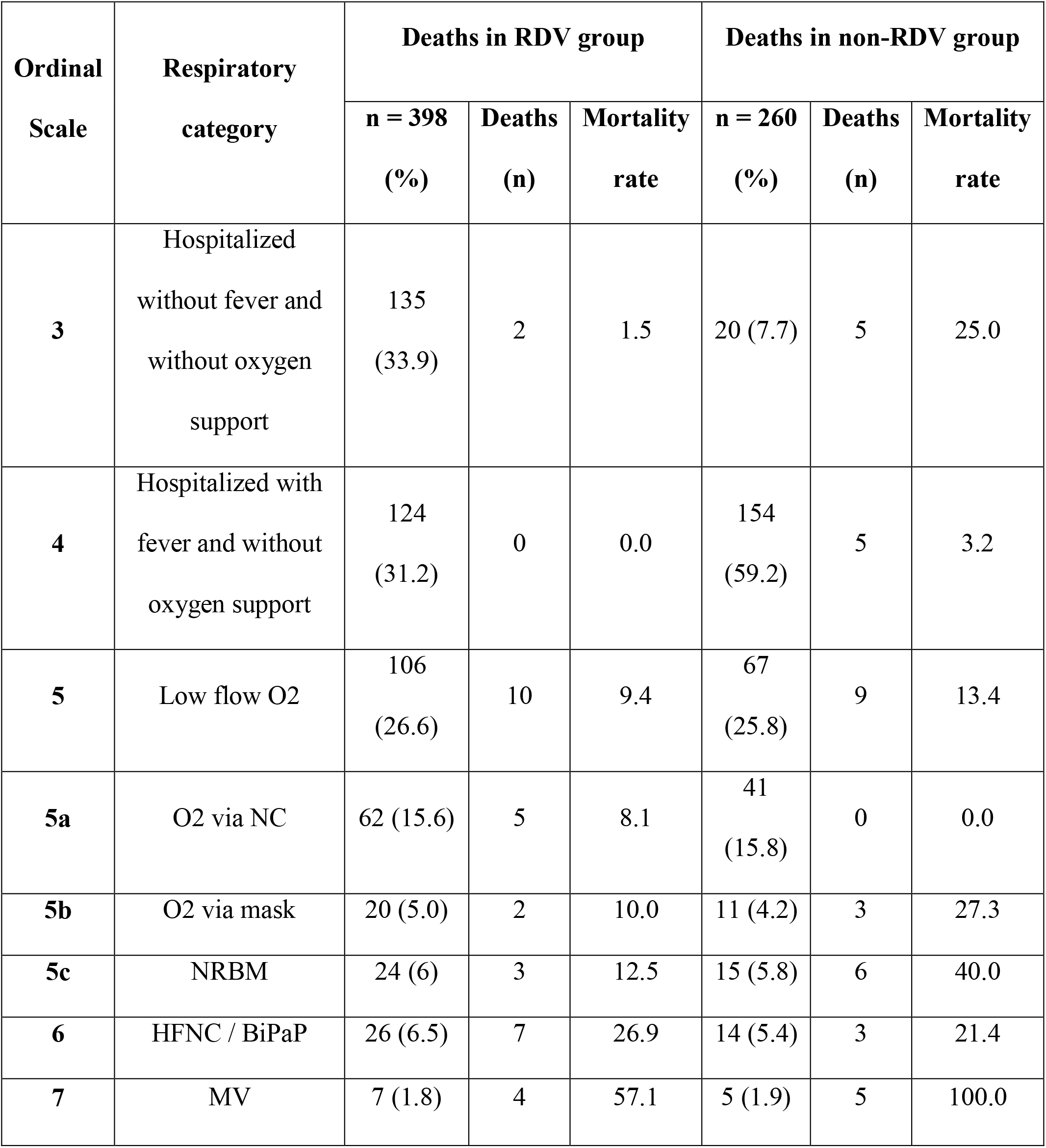
Mortality rate in various respiratory categories

| Ordinal Scale | Respiratory category | Deaths in RDV group |  |  | Deaths in non-RDV group |  |  |
| --- | --- | --- | --- | --- | --- | --- | --- |
|  |  | n = 398 (%) | Deaths (n) | Mortality rate | n = 260 (%) | Deaths (n) | Mortality rate |
| **3** | Hospitalized without fever and without oxygen support | 135 (33.9) | 2 | 1.5 | 20 (7.7) | 5 | 25.0 |
| **4** | Hospitalized with fever and without oxygen support | 124 (31.2) | 0 | 0.0 | 154 (59.2) | 5 | 3.2 |
| **5** | Low flow O2 | 106 (26.6) | 10 | 9.4 | 67 (25.8) | 9 | 13.4 |
| **5a** | O2 via NC | 62 (15.6) | 5 | 8.1 | 41 (15.8) | 0 | 0.0 |
| **5b** | O2 via mask | 20 (5.0) | 2 | 10.0 | 11 (4.2) | 3 | 27.3 |
| **5c** | NRBM | 24 (6) | 3 | 12.5 | 15 (5.8) | 6 | 40.0 |
| **6** | HFNC / BiPaP | 26 (6.5) | 7 | 26.9 | 14 (5.4) | 3 | 21.4 |
| **7** | MV | 7 (1.8) | 4 | 57.1 | 5 (1.9) | 5 | 100.0 |

No significant nehprotoxicity or hepatotoxicity was observed after RDV. Median serum creatinine on day 1, 5 and 7 of RDV administration was 0.9 mg/dl (95% CI 0.9 – 1), 0.8 mg/dl (95% CI 0.8 – 0.9) and 0.9 mg/dl (95% CI 0.9 – 1) respectively. Median alanine transamniase (ALT) on day 1, 5 and 7 of RDV administration was 27.2 U/L, 32.8 U/L and 33.4 U/L respectively. Same values for aspartate transaminase (AST) on day 1, 5 and 7 of RDV administration were 32.7 U/L, 45.9 U/L and 43.6 U/L respectively.

Table 8 and Figure 2 show the cumulative survival probability of patients in both the groups. Figure 2A shows that the difference in probability of the event (death) was statistically not significant (log rank test with p=0.13) for survival in patients in both the groups, though there was a trend towards lower mortality and higher survival in the RDV group.

**Table 8:**
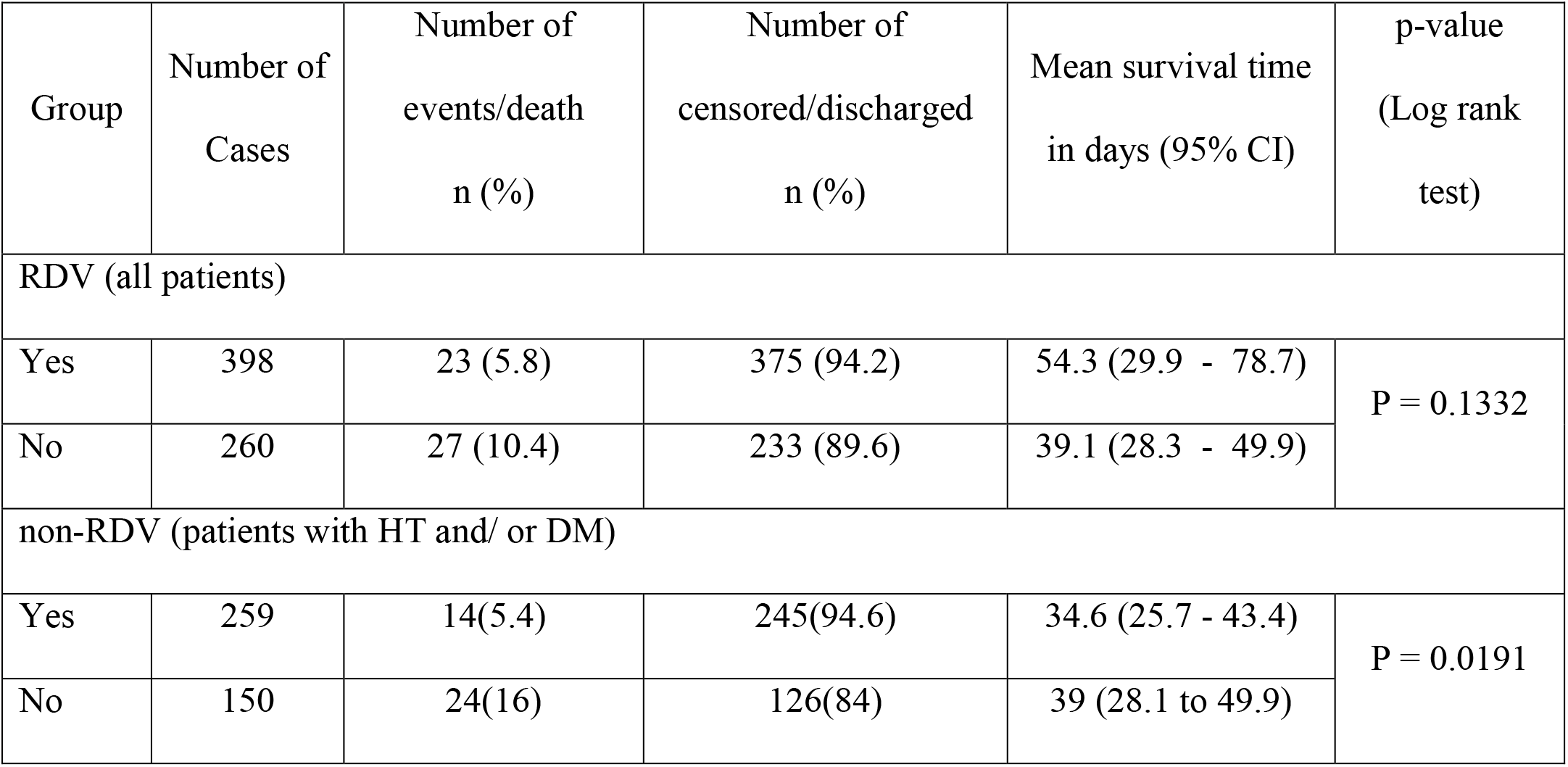
Survival analysis of patients in RDV / non-RDV group; among all patients, and those with HT and/ or DM

**Figure 2A:**
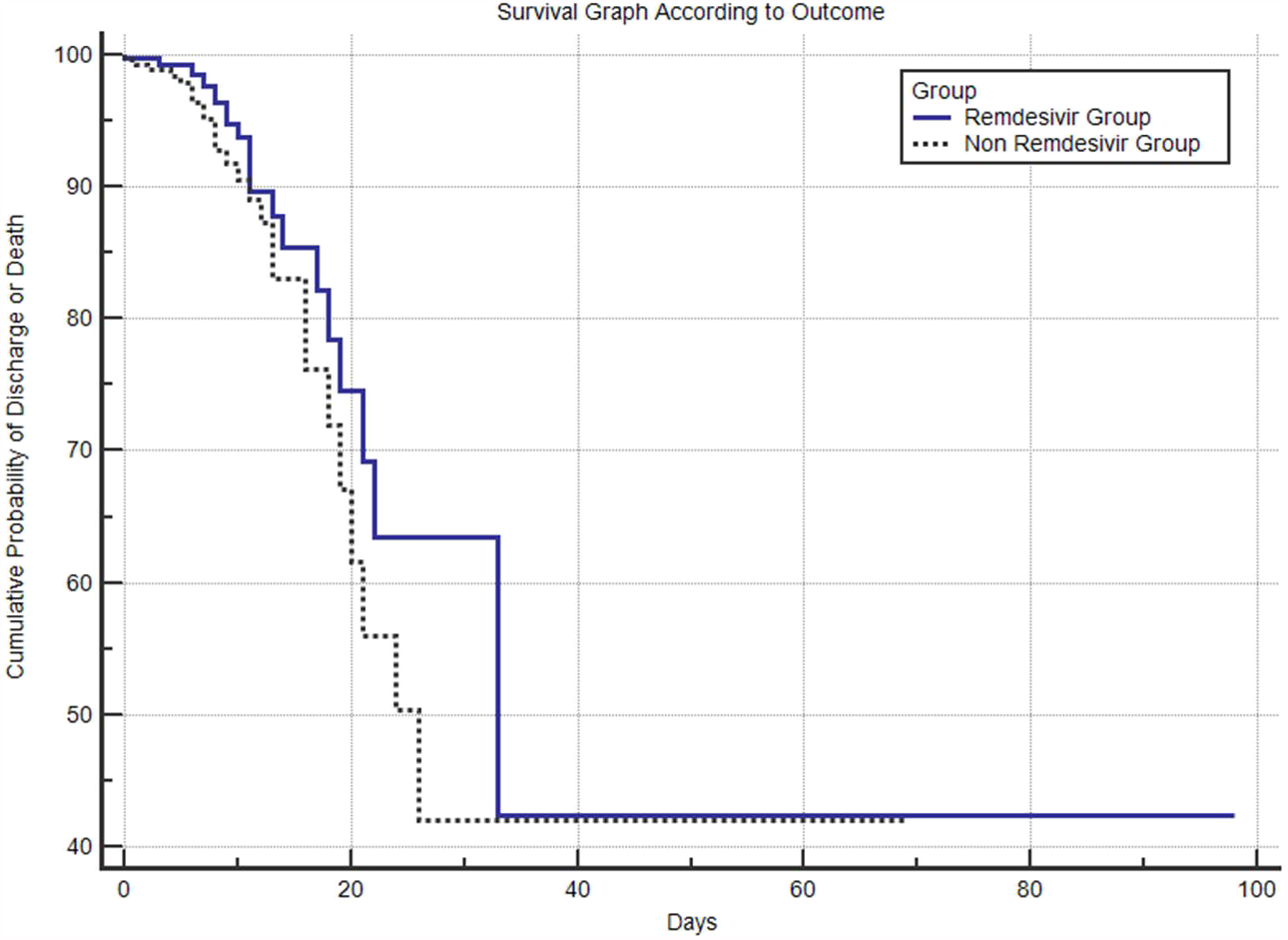
Cumulative probability of discharge or death among all patients in RDV and non-RDV group

**Figure 2B:**
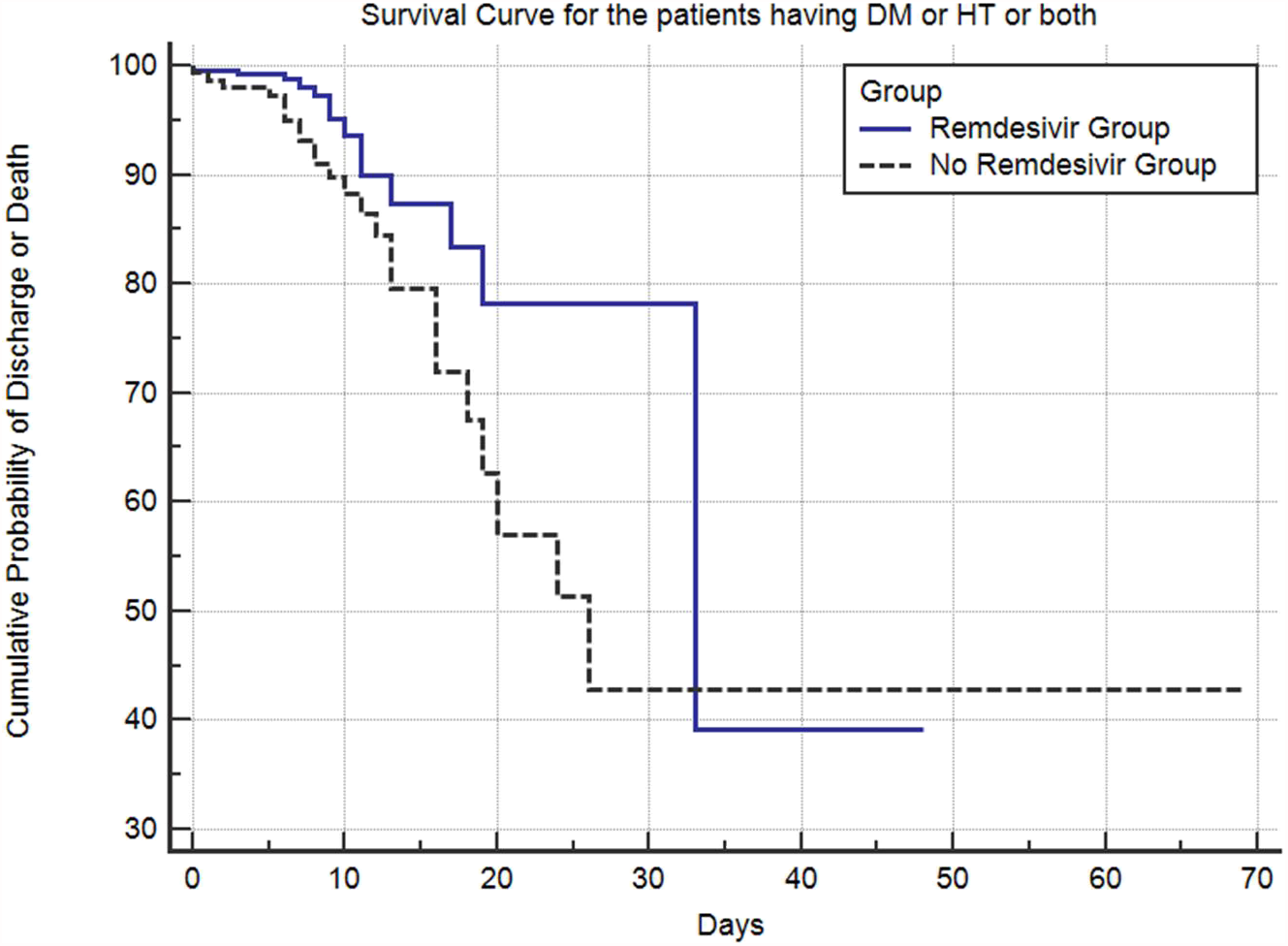
Cumulative probability of discharge or death among patients with HT or/and DM in RDV and non-RDV group

Figure 2B shows survival among patients with HT and/or DM in both the groups; the difference in probability of the event (death) was statistically significant (log rank test with p<0.0191).

## Discussion

This study highlights the role of RDV in the treatment of COVID-19. In this study, the use of RDV was associated with low occurrence of the primary outcomes i.e. death and progression to MV. Mortality in the RDV group was lower. Also, use of early RDV, within 5-10 days of symptoms onset, was associated with lower mortality as compared to late administration of the drug, with mortality being least when RDV was administered within 5 days of symptom onset. Progression to higher ordinal scales suggestive of deteriorating respiratory condition, as well as the mortality across all the ordinal scales (except HFNC/BiPAP) occurred less in the RDV group. This suggests that treatment with RDV may have prevented the progression to more severe disease and probably the role of RDV may exist even in the inflammatory phase of the illness. In our study, 33.9% (n=135) patients in the RDV group had baseline ordinal scale 3 while this proportion was lower in the non-RDV group (7.7%, n=20). This small sample size in the non-RDV group could be the probable reason for high mortality observed (25%).

In the ACTT-1 trial, the largest mortality benefit was seen in patients with the baseline ordinal scale 5 i.e. the patients on low flow oxygen and the benefit of RDV was larger when given earlier in the illness (randomization within 10 days of onset of symptoms) (9). In a study by Oleander et al, RDV was associated with greater recovery and 62% reduced odds of death in severe COVID-19 (10).

Progression to MV in RDV group among ordinal scales 3 to 6 occurred in fewer patients as compared to non-RDV group (4% vs 8.2%). A lower proportion of patients who received RDV while not on oxygen had new need for supplemental oxygen (13.7% vs 22%). In SIMPLE-2 trial, in hospitalized patients with moderate severity (pulmonary infiltrates and room SPO2 >94%, with 83 % patients not receiving oxygen at baseline), patients randomized to a 5 day course of RDV had a statistically significant difference in the clinical status at day 11, but the difference was of uncertain clinical importance. However, the median duration of symptoms in this trial was 8 days before randomization in the RDV group (11).

In the RDV group, the proportion of patients discharged on day 5, day 7 day 10 and day 14 of hospitalization were higher than the non-RDV group. ACTT-1 trial and study by Wang et al showed significantly shorter time to recovery after RDV (9, 12).

In our study, defervescence occurred earlier in the RDV group. Though persistence of fever has not been shown to have direct association with the severity of disease, it may lead to prolonged hospitalization and injudicious use of antibiotics and steroids. In a study by Deborah et al, prolonged fever was associated with hypoxia compared with controls (27.8% vs 0.9%; p < .01). Cases with prolonged fever were also more likely to require ICU admission (11.1% vs 0.9%; p = .05) (13).

Case series have reported safe use in patients with chronic kidney disease and acute kidney injury, with similar observations in our study (14,15).

The interim report of the WHO sponsored SOLIDARITY trial of hospitalized patients with COVID-19 could not demonstrate any mortality benefit with RDV (16). However, according to the data presented by Gilead at the World microbe forum recently, the three retrospective studies including 98,654 patients, showed significantly lower risk for mortality and higher likelihood of discharge by day 28, compared with the matched controls, and reduction in mortality was observed across a spectrum of baseline oxygen requirements (17).

We support the use of RDV early in the course of illness, preferably within 5 days of symptom onset. Although the evidence hitherto favours the use of RDV in hypoxic patients, we support the use of RDV even in absence of hypoxia, especially in patients who have persistent fever and have risk factors for severe disease, like old age and comorbidities.

In the current pandemic situation, when there is a dearth of hospital and ICU beds in resource-limited settings, RDV could shorten the hospital stay, prevent the progression of disease and curtail the need for ICU admission.

## Limitations

Our study has the limitations of a retrospective study. Unequal number of patients and lack of standard classification in the two groups, absence of a standard comparison group, absence of a standard protocol for use of various medications, and missing information, are some of the limitations. Also, use of RDV was done at the discretion of treating clinicians, despite a predefined protocol. The detailed analysis of timing and doses of steroids is not done in this study, which may be an important factor towards the outcome.

## Conclusion

Use of RDV early in the COVID-19 illness is associated with reduced hospital stay and need for supplemental oxygen; and decreases the risk of progression to mechanical ventilation. It may also be associated with decreased mortality in severe as well as mild to moderate category of the disease.

## Data Availability

Data is available in digital form and will be provided if required

## Notes

### Author Contributions

Surabhi Madan and Manish Rana conceived and designed the study and wrote the manuscript. Amit Patel, Bhowmik Meghnathi and Parloop Bhatt approved the final manuscript. Manish Rana and Nirav Bapat performed the analysis. Surabhi Madan, Amit Patel, Kartikae Sharan, Shayon Ghosh, Vishnu Venugopal, Nitesh Shah, Vipul Thakkar, Bhagyesh Shah, Pradip Dabhi, Minesh Patel, Hardik Shah, Rashmi Chovatiya, Maulik Soni, Vineet Sankhla, Vipul Kapoor, Tejas Patel, Kaivan Shah, Ritanshu Chandarana collected and contributed data. All authors read and approved the final manuscript.

## Acknowledgment

The authors thank Dr. Riddhi Parikh, Dr. Disha Patel, Maitri Shah, Sangeetha Arunachalam, Aayushi Singh; Department of Clinical Research, Care Institute of Medical Sciences, Ahmedabad, India, for their assistance in data aggregation and compilation in electronic database.

